# State-wise Economic Burden of Road Traffic Accidents in India

**DOI:** 10.1101/2023.12.21.23300419

**Authors:** S Sajith Kumar, Bhavani Shankara Bagepally, Akhil Sasidharan, Kayala Venkata Jagadeesh, Manickam Ponniah

## Abstract

**Introduction:** Road traffic accidents (RTA) cause multifarious detrimental consequences, including fatality and serious injuries. RTAs cause considerable financial strain on healthcare systems with high costs of medical care, long-term care for the injured, and lost productivity. To assess this burden, we estimated state-wise economic costs of RTA in India. We followed CHEERS Checklist 2022 to present study’s methods and outcomes.

**Methods:** Data were obtained from Global Burden of Disease Injuries and Risk Factors study, Government of India’s budget documents, annual reports of National Health Authority, controller general of accounts, and Economic Surveys. Cost per disability-adjusted life years (DALY), direct costs, indirect costs and total costs associated with RTA burden were estimated for India and by its states.

**Results:** RTA burden declined in all Indian states except Jammu and Kashmir (with a minor increase from 1,458 to 1,564 DALYs per 100,000) and Rajasthan (from 1,275 to 1,394 DALYs per 100,000) during 2010-19 period. Estimated mean cost per DALY in India due to RTA (95%CI) was □1,821(□1,606 to □2,036) [US$ 22(19 to 24)] with a median (IQR) of □1,609(551) [US$ 19 (7)]. As of 2019, the estimated mean total cost of RTA burden in India was □1,017 billion [US$ 12 billion], with a median (IQR) of □731 billion(1,220 billion) [US$ 9 billion(15 billion)].

**Conclusion:** Burden of RTAs declined across the Indian States during 2010-2019, and same was not observed with fiscal implications. The study reiterates the need for strategic road safety interventions to ameliorate socio-economic and health impacts of RTAs in India.

## Introduction

A Road Traffic Accident (RTA) is defined as an incident that takes place on a public road or street, involving at least one moving vehicle, and leading to injuries or fatalities of one or more individuals. (Ruikar 2013) In the context of increasing road mobility, road accidents have become a major concern, particularly in developing countries. (Balakrishnan and Karuppanagounder 2020) With one of the largest road networks in the world, India’s roadways are essential conduits for the movement of people and goods, fostering the growth and development of the country’s economy. (Anon 2021c) However, the increased level of road mobility, coupled with insufficient infrastructure, slack enforcement of traffic laws, and pervasive unsafe driving behaviours, collectively contributes to the increase in road accidents, raising serious issues with public health and safety. (Anon 2022c) The impact of traffic accidents are profound, involving both economic and social implications. RTAs exact a significant human toll, entailing the loss of life, serious injuries, and considerable mental trauma experienced by the affected families.

Given the increasing incidence of RTA and related injuries (Pal *et al*. 2019) along with the substantial regional disparities in the burden they pose, RTAs impose a substantial financial burden on the healthcare system. (Balakrishnan and Karuppanagounder 2020, Ram and Thakur 2022) This is attributable to the high cost of medical care, long-term care for the injured, and lost productivity. (Pal *et al*. 2019, Kovacevic *et al*. 2020) This underscores the need for a comprehensive analysis of the economic implications of the RTA burden across Indian States and these findings can assist policymakers in devising targeted strategies to mitigate and control the RTA burden and its economic consequences. In the absence of comprehensive evidence in the published literature, we aimed to quantitatively estimate the direct and indirect costs of RTAs across various states in India, with a particular emphasis on health expenditure and Years of Life Lost (YLL) attributable to RTA-induced fatalities.

## Methods

### Data Sources

The Institute for Health Metrics and Evaluation (IHME) (Anon 2019a) publishes the Global Burden of Disease (GBD) Injuries and Risk Factors Study, which provides an in-depth estimate of the burden estimates in terms of DALY for India and other countries. (Vos *et al*. 2020) We collected data from the GBD database, encompassing DALY per 100,000 population, DALY per 100,000 population attributable to RTA, and Years of Life Lost (YLL) due to RTA across Indian states over the ten years from 2010 to 2019. We collected state-level healthcare expenditure data from various sources, including budget documents, the Annual Report of the Controller General of Accounts, the Economic Survey, and the National Health Authority’s annual reports. (Anon, 2019b, 2020a, 2021b, a) To address missing data, especially from the northeastern states, we conducted a meticulous compilation of data from these sources across several years. Utilizing a standardized health outcome metric such as DALY per 100,000 population enables comparisons of RTA burden across states. Data on out-of-pocket health expenditure (OOPEs) (Anon 2019b), the value of statistical life in India (Agamoni Majumder 2018), state-wise per capita GDP, per capita GDP in India (Anon 2021a), elasticity coefficient of Willingness to pay (Khan and Mahumud 2015), state-wise consumer price index (Anon 2020b) were systematically collected for subsequent calculations. The health expenditure per capita attributed to RTA was computed by multiplying the percentage of RTA DALY by total DALY by per capita health expenditure. A flowchart illustrating the detailed methodological framework is provided in Figure 1.

**Figure 1:**
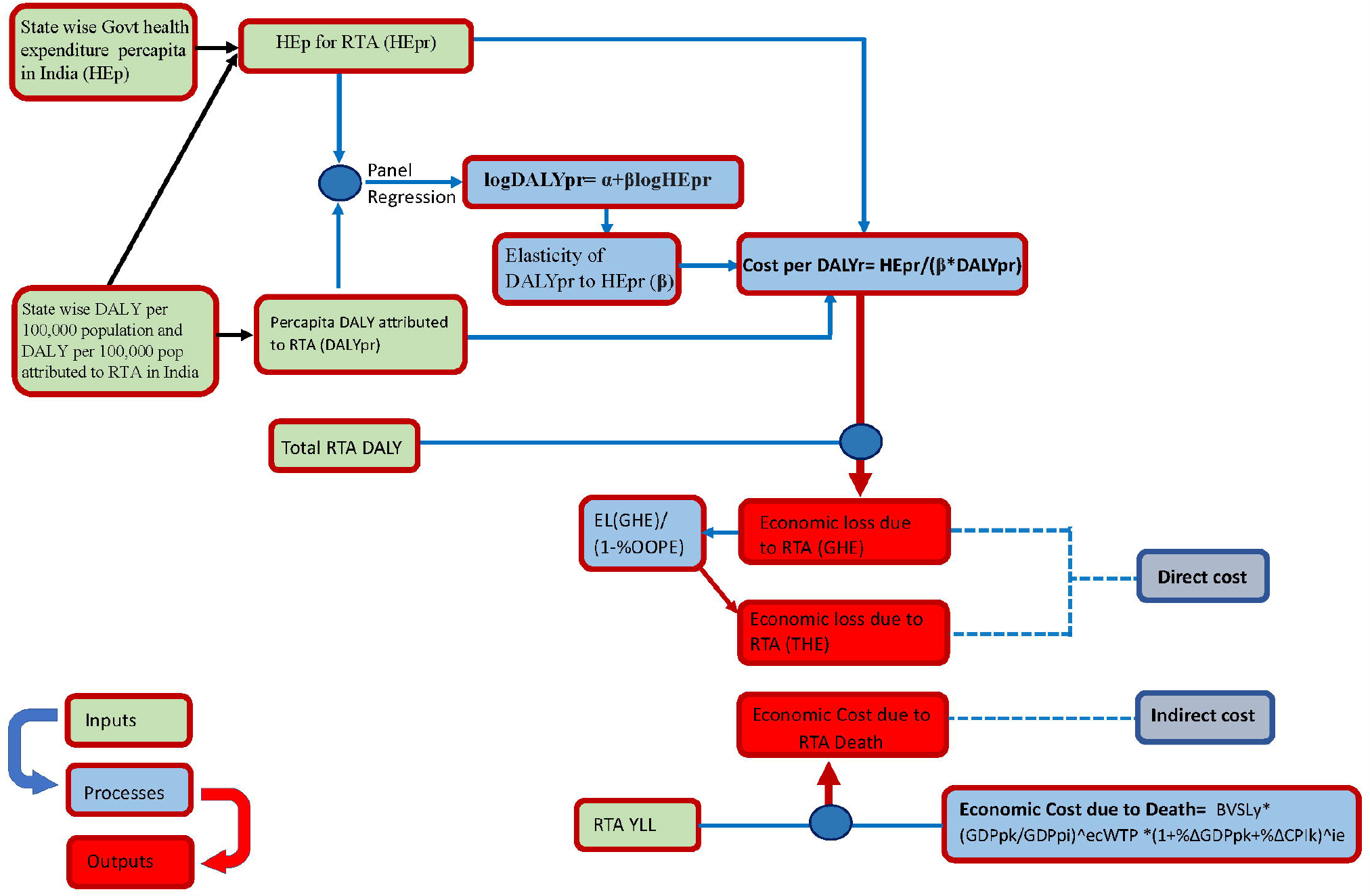
Methodological Framework Flow Chart

### Statistical methods

To estimate state-specific cost per DALY, we employed the methodologies proposed by Daroudi et al. (Daroudi *et al*. 2021) and an adapted approach as recommended by Ochalek et al. (Ochalek *et al*. 2019) To establish the relationship between RTA DALYs per capita and health expenditure per capita attributed to RTA in Indian states, we conducted a panel regression using the following logarithmic regression model:

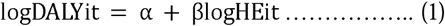

where logDALYit represents the natural logarithm of per capita RTA DALY for state i in year t and logHEit represents the natural logarithm of health expenditure per capita attributed to RTA for state i in year t. The parameter β, the regression coefficient, quantifies the change in RTA DALY associated with a unit change in HEp while holding all other variables constant. The selection between a fixed effects model and a random effects model in panel regression was determined using the Hausman test. The estimated panel regression coefficient (β) was utilized to adjust changes in HEp for the calculation of the cost per RTA DALY for all states.

To calculate the cost per DALY attributed to RTA, we employed the following formula,

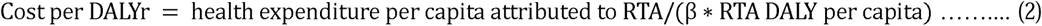

The cost per DALY attributed to RTA functions as a metric for assessing the resources required to attain an additional healthy year of life within a population. Further, to estimate the economic cost of RTA burden from a government health expenditure (GHE) perspective, we used the following formula,

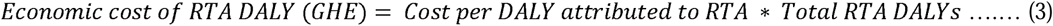

Similarly, for the estimation of the total economic cost of RTA DALYs encompassing both government and private spending, we acquired data on the proportion of OOPEs. This information was then integrated as an adjustment factor for calculation, as outlined in the references (Anon 2019b, 2022a). The formula used to calculate the total economic cost of RTA burden considering both GHE and private spending’ is, Ec(GHE)/1-Percentage of OOPE, where Ec(GHE) is the economic cost of RTA burden from the government perspective. In cases where state specific OOPEs data were unavailable, we opted for using national averages.

Additionally, we estimated the economic cost of mortality burden within individual States in India, employing a methodology derived from Balakrishnan S et al. (2020) that assessed the economic cost associated with cause-specific PM2.5 mortality (Balakrishnan and Karuppanagounder 2020). The economic cost of mortality is calculated using the formula,

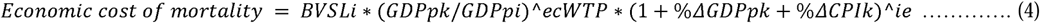

Where BVSL*i* is the base year value of statistical life in India, GDPpk is per capita GDP in state *k*, GDPp*i* is per capita GDP in India, *EcWTP* is the elasticity coefficient of WTP, CP*Ik* is consumer price index for state *k* and *ie* is income elasticity for health.

Further, the economic cost of the RTA burden is obtained by multiplying the Economic cost of mortality with YLL due to RTA.

### Data analysis

All analyses were performed using Stata V.17(StataCorp. 2021). Values are reported as the mean with a 95% confidence interval (CI) and the median with the interquartile range (IQR). We used graphical representations to further visualise the trends and economic costs of RTA burden among individual states in India. We have used crude DALY to account for the population difference across different states in India and further updated the model and analysis using the latest available Indian data. Considering the potential impact of the COVID-19 pandemic on disease burden and gross productivity, and data constraints, this study focused solely on a more recent pre-pandemic period, 2010 to 2019. We adhered to the CHEERS Checklist 2022 to ensure a concise and thorough presentation of our study’s methodology and findings and included the CHEERS checklist as appendix 1 in the supplementary material. All the estimated numbers represent a single year; therefore, the conversion factor for a single year (1 US$ = □82.4) is applied to report the values in USD from Indian rupees (□). (Anon 2022b)

## Results

### Trends in RTA burden in India

We analyzed the trends in RTA burden per 100,000 population in India from 2010 to 2019. During this period, the disease burden (as measured using DALY) due to RTA declined in all Indian states except Jammu and Kashmir, where it exhibited a minor increase from 1,458 to 1,564 DALYs, and in Rajasthan, where it rose from 1,275 to 1,394 DALYs (Figure 2, Supp Fig 1). The data for 2019 shows the states with the lowest percent of RTA burden compared to the total disease burden include Meghalaya (0.0222%), Bihar (0.0255%), Assam (0.0260%), and Nagaland (0.0279%). On the other hand, states with the highest share of RTA burden as a percentage of total disease burden are Punjab (0.0411%), Haryana (0.0499%), Uttarakhand (0.0505%), and Jammu & Kashmir (0.0525%) (Supp Figure 2).

**Figure 2:**
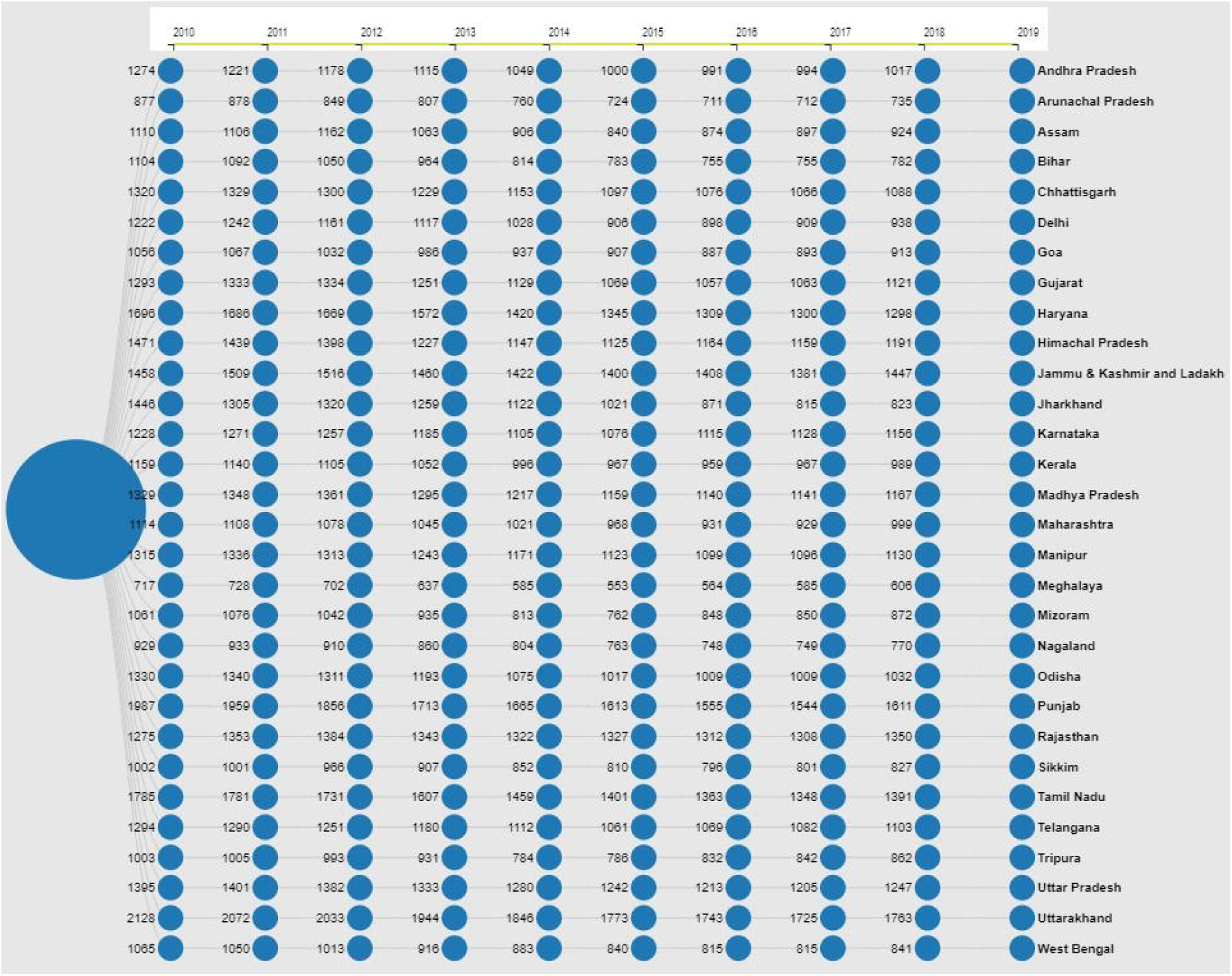
Road traffic accidents DALY per 100,000 population among Indian States

### Cost per DALYs due to RTA by States

The estimated mean cost per DALY due to RTA burden (95% CI) was □1,821 (□1,606 to □2,036) [US$ 22 (19 to 24)], with a median (IQR) of □1,609 (551) [US$ 19(7)]. States such as Arunachal Pradesh (□3,234) [US$ 38], Meghalaya (□2,991) [US$ 36], Sikkim (□2,848) [US$ 34], followed by Nagaland (□2,631) [US$ 31] exhibit higher cost per DALY due to RTA burden. Conversely, Uttar Pradesh (□1,054) [US$ 13], Punjab (□1,143) [US$ 14], and Uttarakhand (□1,159) [US$ 14] report the lowest estimates (Supp Fig 3).

### Direct cost of RTA from government perspective by States

The estimated mean direct cost of RTA burden in India, considering the GHE perspective for the year 2019, was □743 million [US$ 9 million], with a median (IQR) of □567 million (1,060 million) [US$ 7 million (13 million)]. States with the highest direct costs due to RTA burden include Uttar Pradesh at □3,224 million [US$ 38 million], followed by Maharashtra at □2,058 million [US$ 24 million], and West Bengal at □1,560 million [US$ 19 million]. Conversely, states with relatively lower direct costs due to RTA burden are Sikkim at □16 million [US$ 0.19 million], Mizoram at □29 million [US$ 0.35 million], Goa at □37 million [US$ 0.44 million], Nagaland at □40 million [US$ 0.48 million], and Arunachal Pradesh at □41 million [US$ 0.49 million]. (Supp Fig 4).

### Total direct cost of RTA by States

The estimated mean direct total cost of RTA burden in India in 2019 was □1,534 million [US$ 18 million] with a median (IQR) of □1,155 million (2,588 million) [US$ 13 million (31 million)]. Among Indian states, Uttar Pradesh reported the highest direct total cost (□6,200 million) [US$ 74 million], followed by Maharashtra (□3,957 million) [US$ 47 million], Gujarat (□3,632 million) [US$ 43 million], and West Bengal (□3,000 million) [US$ 36 million] for RTA burden. In contrast, States like Sikkim (□55 million) [US$ 0.65 million], Goa (□58 million) [US$ 0.69 million], Nagaland (□73 million) [US$ 0.87 million], and Arunachal Pradesh (□74 million) [US$ 0.88 million] showed lower total direct costs compared to other States. (Fig 3)

**Figure 3:**
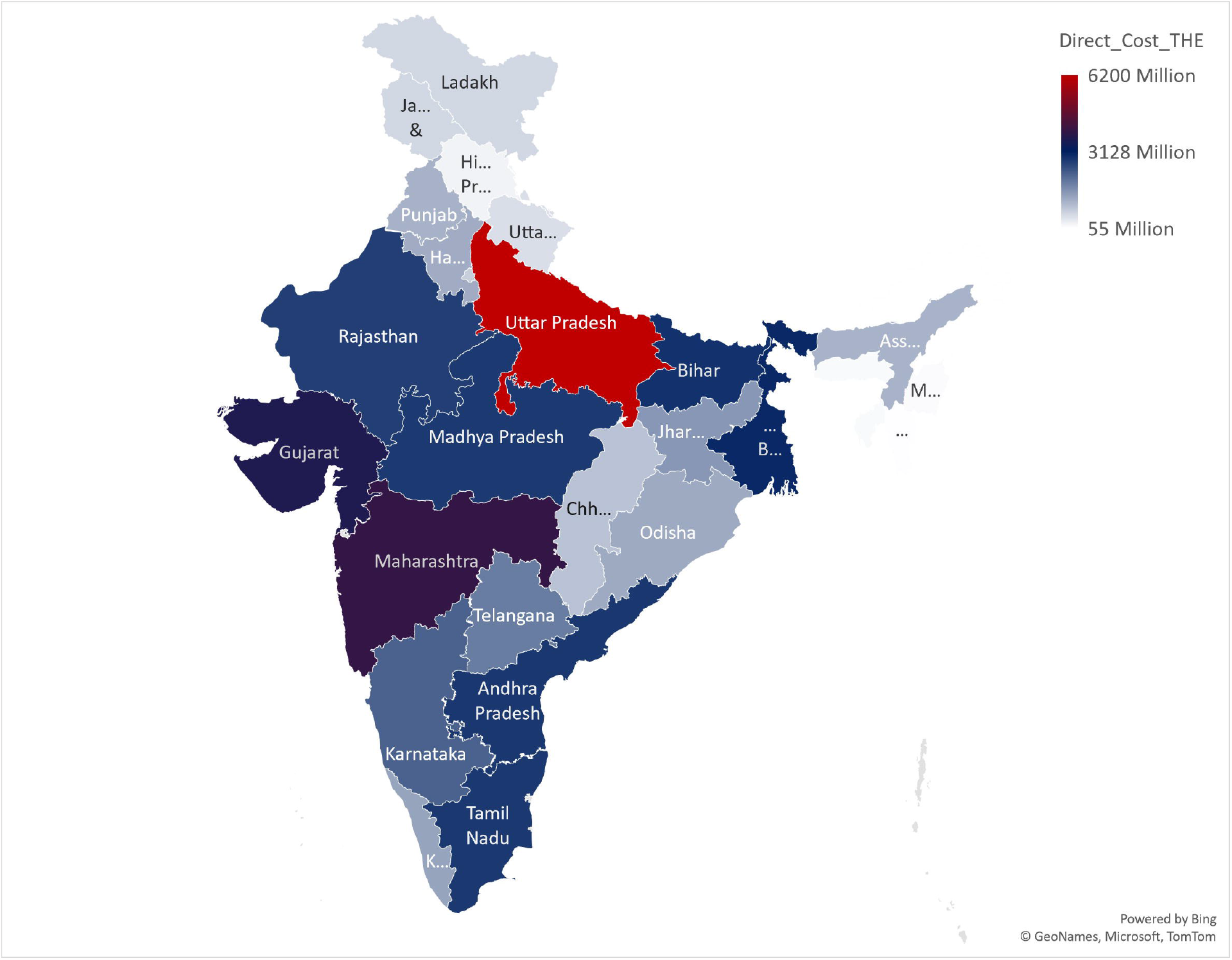
Direct cost of RTA in Indian states.

### Indirect costs of RTA by States

The estimated mean indirect costs of RTA burden in India in 2019 was □1,015 billion [US$ 12 billion] with a median (IQR) of □729 billion (1,221 billion) [US$ 9 billion (15 billion)]. When considering the indirect costs due to the RTA burden, it becomes evident that states like Punjab (3,476 billion) [US$ 42 billion], Arunachal Pradesh (3,461 billion) [US$ 42 billion], Sikkim (3,177 billion) [US$ 39 billion], and Himachal Pradesh (2,643 billion) [US$ 32 billion] report higher costs. In contrast, states such as Maharashtra (22 billion) [US$ 0.27 billion], Bihar (23 billion) [US$ 0.28 billion], West Bengal (23 billion) [US$ 0.28 billion], and Uttar Pradesh (28 billion) [US$ 0.34 billion] exhibit comparatively lower indirect costs. (Supp Fig 5)

### Total costs of RTA by States

In 2019, the estimated mean total costs of RTA burden in India was □1,017 billion, [US$ 12 billion] with a median (IQR) of □731 billion (1,220 billion) [US$ 9 billion (15 billion)]. Among Indian states, Punjab had the highest cost due to RTA burden, reaching □3,477 billion [US$ 42 billion], followed by Arunachal Pradesh (□3,461 billion) [US$ 42 billion], Sikkim (□3,177 billion) [US$ 39 billion], and Himachal Pradesh (□2,643 billion) [US$ 32 billion]. Conversely, states like Maharashtra, Bihar, West Bengal (all □26 billion) [US$ 0.32 billion], and Uttar Pradesh (□34 billion) [US$ 0.41 billion] had comparatively lower indirect costs. (Supp Fig 6)

## Discussion

We examined trends in population adjusted RTA in India from 2010 to 2019 and documented a decline in the RTA burden in majority of the Indian States. On the other hand, the financial implication of the RTA burden was highly variable across States and did not decline over time.

The observed decline in RTA burden from 2010 to 2019 across the States suggests a positive trend in road safety measures and interventions during this period. Notably, Meghalaya had the lowest RTA burden, and in contrast, Jammu & Kashmir, Uttarakhand, and Punjab face a more pronounced challenge related to the RTA burden.

The cost per DALY is a crucial indicator of the economic impact of RTA-related health conditions, representing the costs incurred for each DALY lost due to RTA. The cost per DALY attributed to the RTA burden exhibits notable variation across Indian States. The States from the northeastern region including Arunachal Pradesh, Meghalaya followed by Sikkim report significantly higher costs, suggesting a greater financial burden associated with RTA. The observed phenomenon of the lowest RTA burden coupled with a higher cost per DALY in northeastern States may be influenced by unique geographical and infrastructural challenges specific to the northeastern region. In contrast, States like Uttar Pradesh, Punjab, and Uttarakhand exhibit the lowest cost per DALY due to the RTA burden. Disparities in healthcare infrastructure, accessibility, and response capabilities can further contribute to elevated costs per DALY despite a lower overall disease burden. However, a comprehensive analysis would be needed to assess the specific interventions, healthcare infrastructure, and preventive measures contributing to this efficiency in States with lower costs per DALY.

The elevated direct costs attributed to RTAs in Uttar Pradesh followed by Maharashtra and West Bengal may be influenced by factors such as population density, traffic conditions, and the availability of healthcare resources and services. These factors contribute to the increased financial burden associated with managing the impact of RTAs. The study conducted by Ram et al. also noted a significant interstate variation in traffic accidents, both in terms of prevalence and associated OOPEs in India. (Ram and Thakur 2022) Interestingly, the highest expenditure is observed in States with lower prevalence, such as Uttar Pradesh and Jharkhand, while the lowest amount is found in the state of Goa. (Ram and Thakur 2022) The higher indirect costs show the strain on both the affected individuals/households and the broader economy, emphasizing the need for preventive measures and interventions to reduce the overall economic burden of RTAs.

The overall economic impact of the RTA burden in India also exhibited a considerable variation across the states. Punjab has the highest economic burden from RTAs, followed closely by Arunachal Pradesh, and Sikkim. Conversely, states like Maharashtra, Bihar, and West Bengal had relatively lowest total costs due to RTA. Similar results were found in 2005, where RTAs caused 110,000 deaths, 2.5 million hospitalizations, and economic losses equivalent to 3% of the GDP, revealing alarming statistics on fatalities, hospitalizations, and economic repercussions. (Gururaj 2008) These results provide valuable insights into the financial implications of the RTA burden in India, with certain States experiencing a considerably higher economic burden compared to others.

Understanding interstate variations can help policymakers to develop targeted strategies and interventions to manage and reduce the economic burden associated with RTAs in different regions, ultimately improving road safety and minimizing the economic consequences of accidents. Given the substantial economic burden linked to RTAs, the key emphasis should be on preventive measures and reducing fatalities resulting from RTAs. Strategies collectively involving public engagement for creating awareness to promote safe road behavior (Hamann *et al*. 2021), technological interventions to enhance road safety through systems like advanced driver assistance (Eskandari Torbaghan *et al*. 2022), legal measures enforcing traffic regulations and penalties (Verma *et al*. 2011), and infrastructural improvements focusing on developing safer road designs and traffic management (Wiethoff *et al*. 2012) contribute to mitigating the economic impact of RTAs and fostering safer road environments. A two-year pilot study on Speed-camera networks has revealed that the costs of camera enforcement were nearly one-third of the total costs of prevented casualties. (Gains *et al*. 2004) In the United Kingdom, speed cameras yielded an average return of 5 times the investment after 1 year and 25 times after 5 years. In 2003, benefits from avoided injuries exceeded £221 million, more than 4 times the cost of enforcement. (Gains *et al*. 2004, Richter *et al*. 2006, Goniewicz *et al*. 2015) Some Indian states, like Tamil Nadu, have already established a special taskforce on road safety composed of experts to enhance overall safety on roadways. As part of the government’s new initiative, *‘Nammai Kakkum 48’* (saving our lives in 48 hours), they also provide free emergency treatment to RTA victims for two days. (Anon 2021d)

The study by Antony, 2021 found that the mean time for RTA victims to reach any healthcare facility was 3 hours, with 55% reaching within the golden hour. Among those beyond the golden hour, delays were attributed to factors such as unavailability of transporting vehicles, communication issues, prolonged travel, lack of knowledge about nearby facilities, nonavailability of attenders and financial constraints. (Antony *et al*. 2021) The variation in cost per DALY and economic costs of RTA burden across states underscores the disparities in the economic impact of RTA-related diseases and highlights the need for tailored interventions to address the issue effectively. Legislation can be an effective instrument for reducing unsafe behaviours among road users and thereby improving road safety. Setting up speed limits for densely populated regions, restricting alcohol levels in drivers, imposing fines, and mandating the use of protective helmets/seat belts are all important legislative measures which have been proven to reduce RTAs. (Anon 2018) Within the health system, there is a pressing need to strengthen emergency response systems and trauma care facilities to ensure swift and efficient medical assistance for RTA victims. Additionally, the establishment of comprehensive rehabilitation services is imperative to support the recovery and reintegration of individuals affected by RTAs. Public awareness campaigns play a pivotal role in promoting road safety by educating the public about preventive measures and emphasizing the importance of using protective gear. Advocacy for robust Road Safety Policies is essential to create a supportive framework for effective prevention and management of RTAs. Furthermore, ensuring the availability of emergency life-saving medicines and blood for RTAs and adequately trained staff is paramount within the health system.

## Limitations

Our study had few limitations. Firstly the use of publicly available secondary data of various types may have its implications on definitions used and methods employed by each of the agencies while putting together data. Hence, when projecting our findings beyond the other settings, one must examine the specificities of our data sources and methodology. Secondly, we believe that we could have underestimated the health expenditure attributable to RTA since state-specific data on healthcare spending exclusive for RTA burden was not available. Thirdly, given the uniqueness of each setting, population, and healthcare system, generalizing our findings to other scenarios requires caution.

## Conclusions and recommendations

On the basis of publicly available data, we documented considerable decline in RTA burden over a decade. Nevertheless, we observed substantial state-wise differences in cost per DALY and economic costs. These findings underscore the urgency for targeted interventions, especially in states with the highest economic impact, to mitigate the burden of RTAs. Additionally, states with lower economic burdens can serve as models for effective road safety measures, offering valuable insights for broader implementation.

## Data Availability

All data produced in the present study are available upon reasonable request to the authors

## Acknowledgements

None

