## Supplementary Materials for "State-wise Economic Burden of Road Traffic Accidents in India"

**Supplementary Figures: Page 2-7**

Supplementary Figure 1- Change in RTA DALY burden in India	2

Supplementary Figure 2- Change in RTA DALY burden proportion to total DALY in India	3

Supplementary Figure 3- State wise cost per DALY due to RTA burden	3

Supplementary Figure 4- Direct cost associated with RTA burden (Government health expenditure)	4

Supplementary Figure 5- Indirect cost associated with RTA burden	5

Supplementary Figure 6- Total economic cost associated with RTA burden	6

CHEERS 2022 Checklist	8

Supplementary Figure 1- Change in RTA DALY burden in India

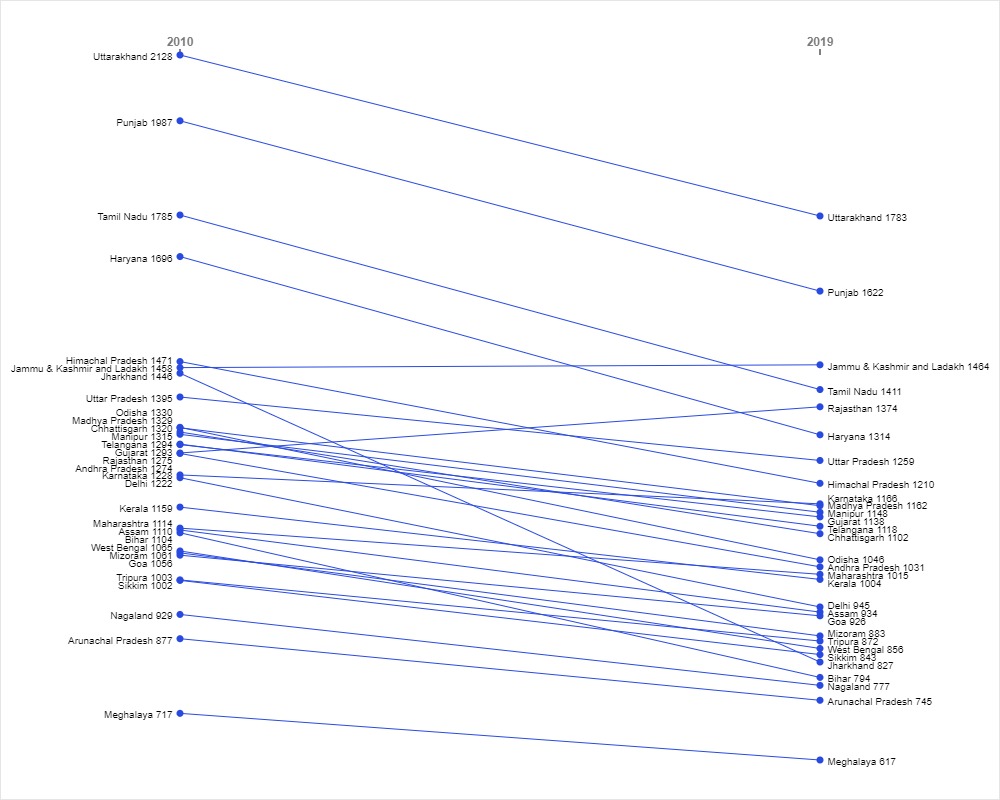

Supplementary Figure 2- Change in RTA DALY burden proportion to total DALY in India

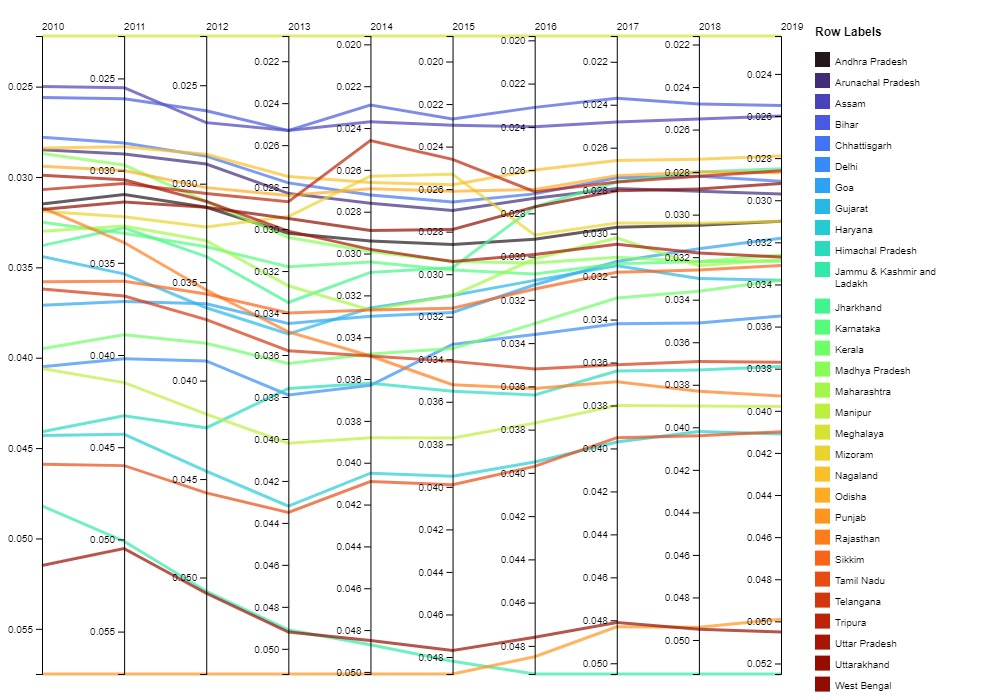

Supplementary Figure 3- State wise cost per DALY due to RTA burden

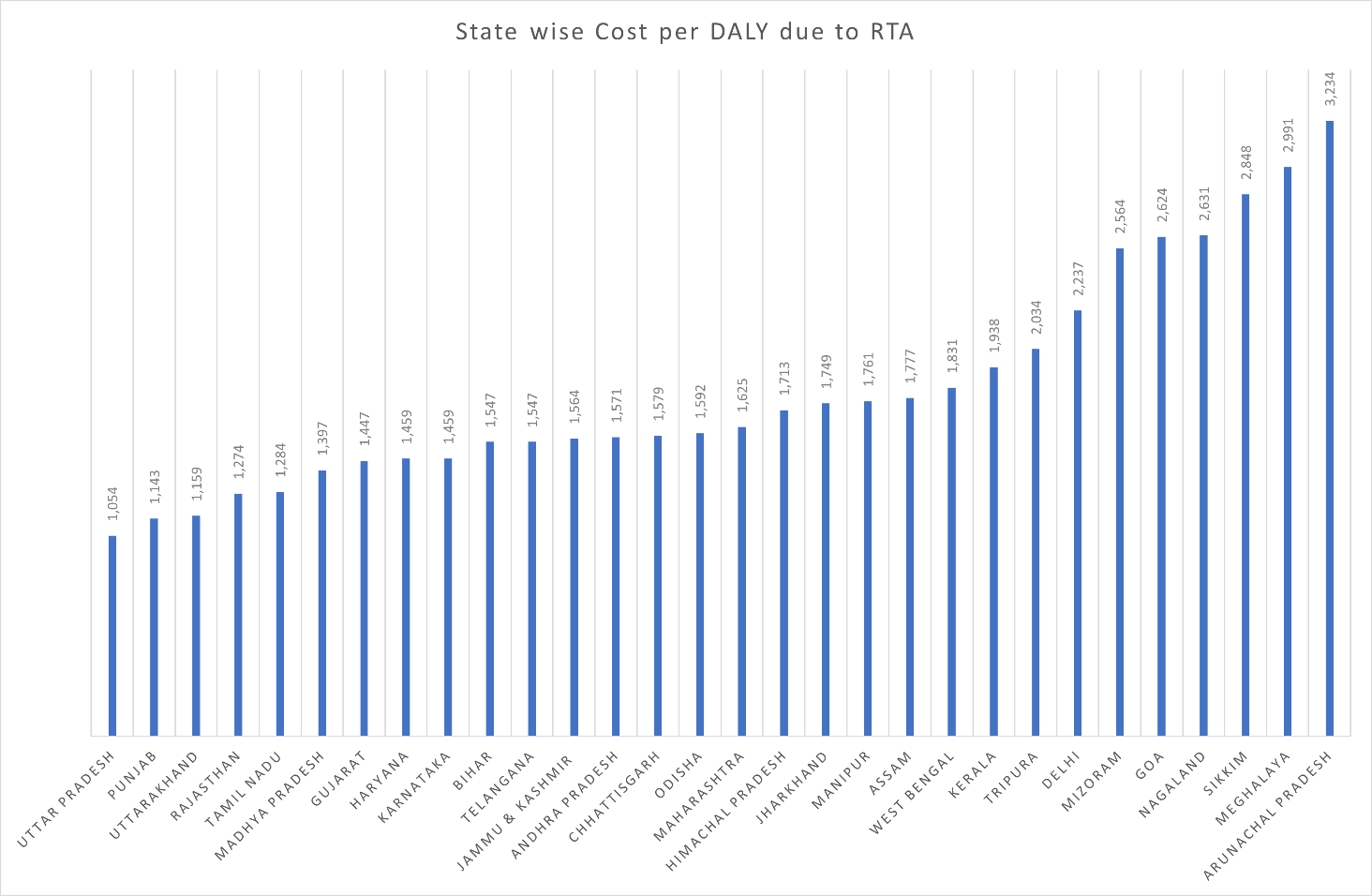

Supplementary Figure 4- Direct cost associated with RTA burden (Government health expenditure)

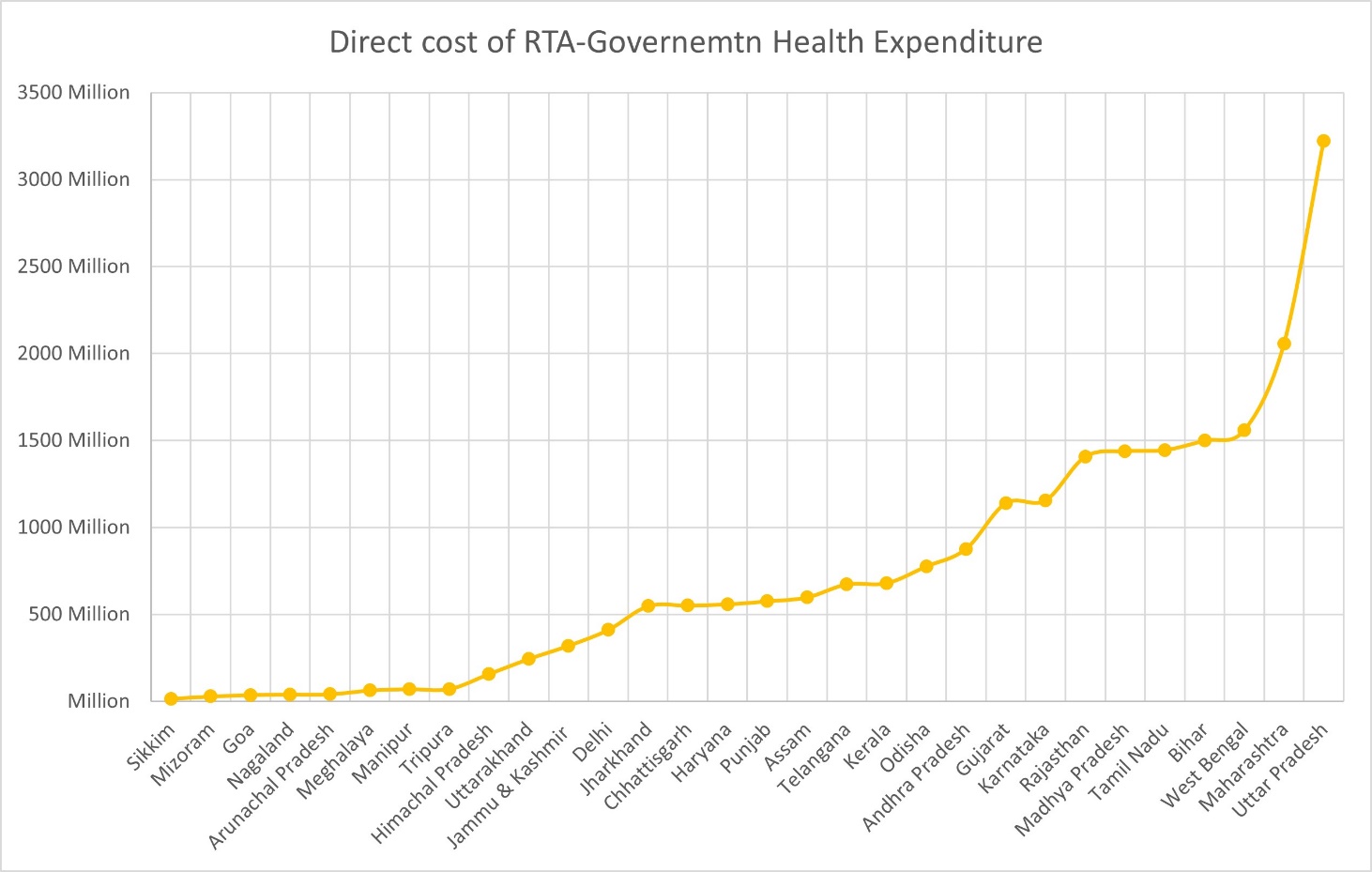

Supplementary Figure 5- Indirect cost associated with RTA burden

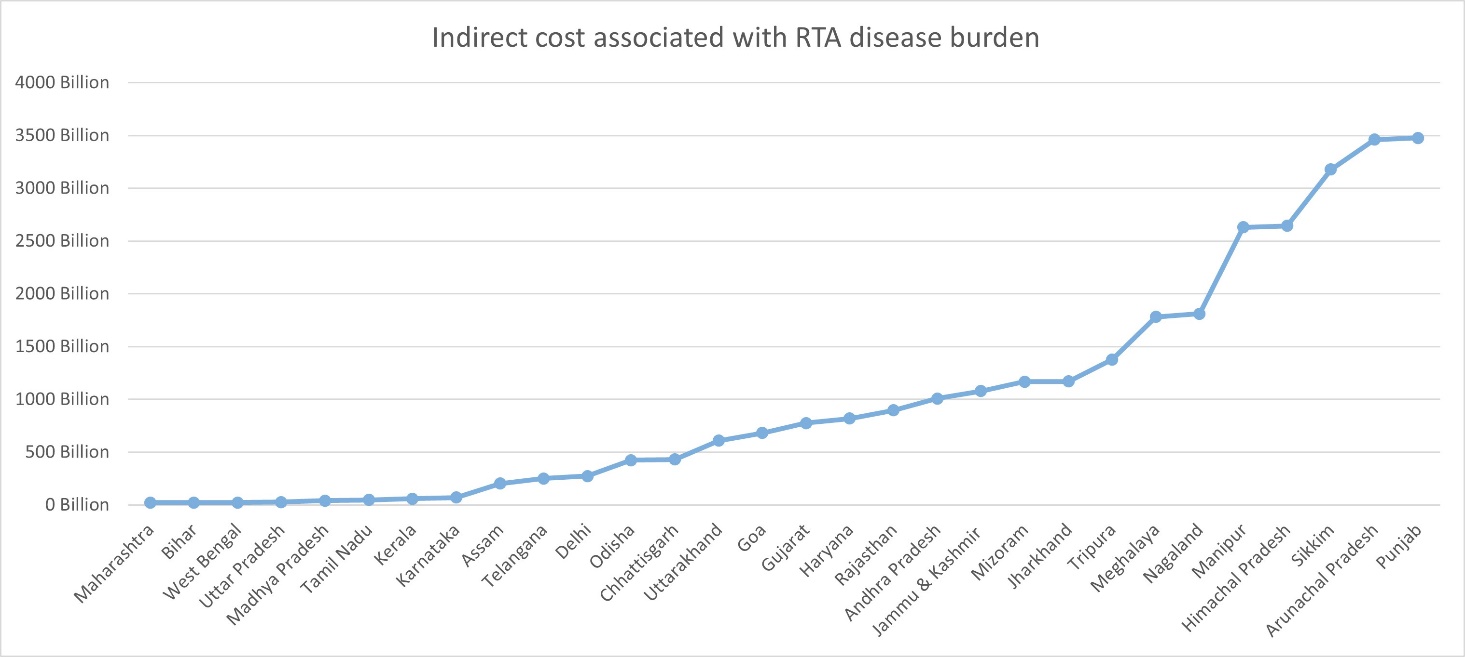

Supplementary Figure 6- Total economic cost associated with RTA burden

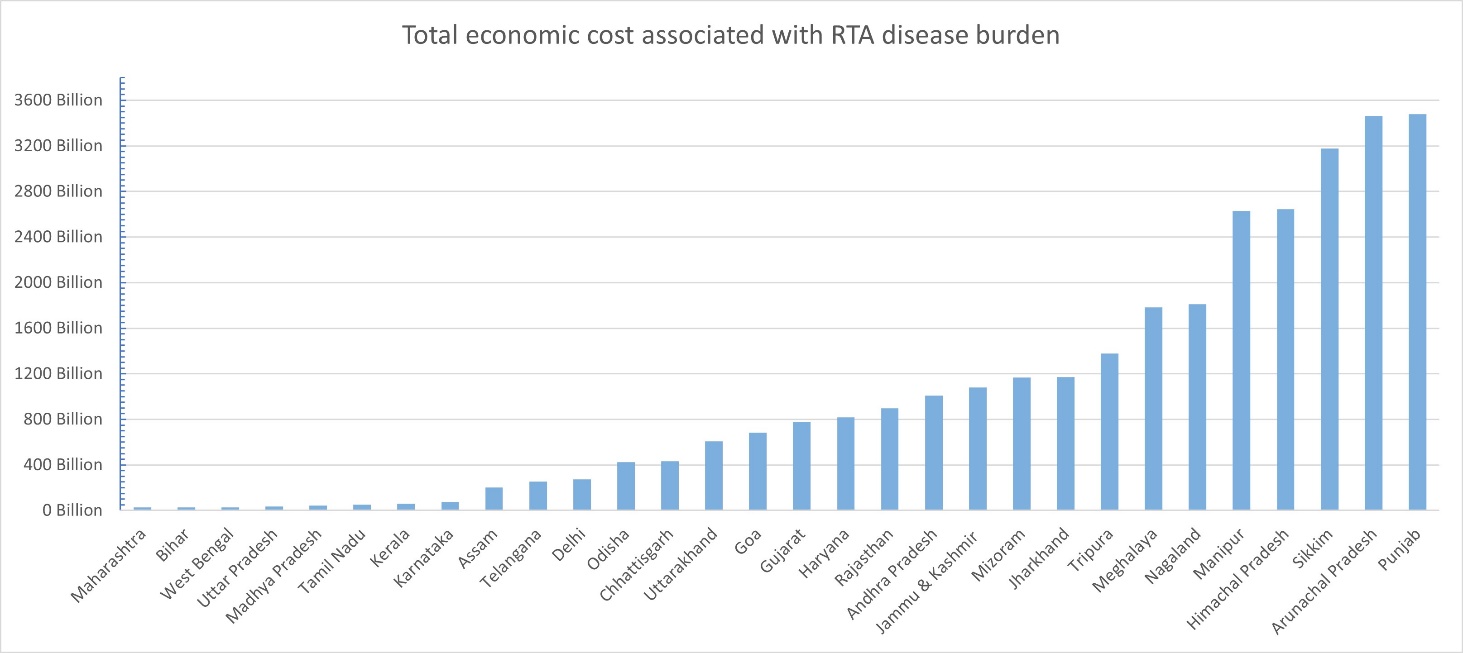

CHEERS 2022 Checklist

| **Topic** | **No.** | **Item** | **Location where item is reported** |
| --- | --- | --- | --- |
| **Title :** State-wise Economic Burden of Road Traffic Accidents in India. | | | |
|  | 1 | Identify the study as an economic evaluation and specify the interventions being compared. | NA |
| **Abstract** | | | |
|  | 2 | Provide a structured summary that highlights context, key methods, results, and alternative analyses. | Page 1, lines 2-22 |
| **Introduction** |  |  |  |
| **Background and objectives** | 3 | Give the context for the study, the study question, and its practical relevance for decision making in policy or practice. | Page 2, Lines 27-48 |
| **Methods** | | | |
| **Health economic analysis plan** | 4 | Indicate whether a health economic analysis plan was developed and where available. | NA |
| **Study population** | 5 | Describe characteristics of the study population (such as age range, demographics, socioeconomic, or clinical characteristics). | NR |
| **Setting and location** | 6 | Provide relevant contextual information that may influence findings. | Page 3-4, lines 52-106 |
| **Comparators** | 7 | Describe the interventions or strategies being compared and why chosen. | NA |
| **Perspective** | 8 | State the perspective(s) adopted by the study and why chosen. | Page 3-4, Lines 70-106 |
| **Time horizon** | 9 | State the time horizon for the study and why appropriate. | Page 3, lines 52-69 |
| **Discount rate** | 10 | Report the discount rate(s) and reason chosen. | NA |
| **Selection of outcomes** | 11 | Describe what outcomes were used as the measure(s) of benefit(s) and harm(s). | Page 4, lines 105-122 |
| **Measurement of outcomes** | 12 | Describe how outcomes used to capture benefit(s) and harm(s) were measured. | Page 3-4, lines 70-106 |
| **Valuation of outcomes** | 13 | Describe the population and methods used to measure and value outcomes. | Page 3-4, lines 70-106 |
| **Measurement and valuation of resources and costs** | 14 | Describe how costs were valued. | NA |
| **Currency, price date, and conversion** | 15 | Report the dates of the estimated resource quantities and unit costs, plus the currency and year of conversion. | Page 5, lines 116-118 |
| **Rationale and description of model** | 16 | If modelling is used, describe in detail and why used. Report if the model is publicly available and where it can be accessed. | NA |
| **Analytics and assumptions** | 17 | Describe any methods for analysing or statistically transforming data, any extrapolation methods, and approaches for validating any model used. | NA |
| **Characterising heterogeneity** | 18 | Describe any methods used for estimating how the results of the study vary for subgroups. | NA |
| **Characterising distributional effects** | 19 | Describe how impacts are distributed across different individuals or adjustments made to reflect priority populations. | NA |
| **Characterising uncertainty** | 20 | Describe methods to characterise any sources of uncertainty in the analysis. | NR |
| **Approach to engagement with patients and others affected by the study** | 21 | Describe any approaches to engage patients or service recipients, the general public, communities, or stakeholders (such as clinicians or payers) in the design of the study. | NA |
| **Results** | | | |
| **Study parameters** | 22 | Report all analytic inputs (such as values, ranges, references) including uncertainty or distributional assumptions. | Page 5-7 Lines, lines 120-171 |
| **Summary of main results** | 23 | Report the mean values for the main categories of costs and outcomes of interest and summarise them in the most appropriate overall measure. | Page 7-8, lines 173-192 |
| **Effect of uncertainty** | 24 | Describe how uncertainty about analytic judgments, inputs, or projections affect findings. Report the effect of choice of discount rate and time horizon, if applicable. | NR |
| **Effect of engagement with patients and others affected by the study** | 25 | Report on any difference patient/service recipient, general public, community, or stakeholder involvement made to the approach or findings of the study | NA |
| **Discussion** | | | |
| **Study findings, limitations, generalisability, and current knowledge** | 26 | Report key findings, limitations, ethical or equity considerations not captured, and how these could affect patients, policy, or practice. | Page 7-11, lines 173-265 |
| **Other relevant information** | | | |
| **Source of funding** | 27 | Describe how the study was funded and any role of the funder in the identification, design, conduct, and reporting of the analysis | Page 11, lines 267-269 |
| **Conflicts of interest** | 28 | Report authors conflicts of interest according to journal or International Committee of Medical Journal Editors requirements. | Page 11, lines 270 |

*NA- Not applicable, NR- Not reported*

*Source:* Husereau D, Drummond M, Augustovski F, et al. Consolidated Health Economic Evaluation Reporting Standards 2022 (CHEERS 2022) Explanation and Elaboration: A Report of the ISPOR CHEERS II Good Practices Task Force. Value Health 2022;25. <doi:10.1016/j.jval.2021.10.008>
